# Modest effects of contact reduction measures on the reproduction number of SARS-CoV-2 in the most affected European countries and the US

**DOI:** 10.1101/2020.04.20.20067538

**Authors:** Armin Ensser, Pia Überla, Klaus Überla

## Abstract

Population density, behaviour and cultural habits strongly influence the spread of pathogens. Consequently, key epidemiological parameters may vary from country to country. Many estimates of SARS-CoV-2 and COVID-19 strongly depend on testing frequency and case definitions. The fatal cases due to SARS-CoV2 could be a more reliable parameter, since missing of deaths is less likely. We analysed the dynamics of new infection and death cases to estimate the daily reproduction numbers (R_t_) and the effectiveness of control measures in the most affected European Countries and the US. In summary, calculating R_t_ based on the daily number of deaths as well as of new infections may lead to more reliable estimates than those based on infection cases alone, as death based R_t_ are expected to be less susceptible to testing bias or limited capacities.

---

Population density, behaviour and cultural habits strongly influence the spread of pathogens. Consequently, key epidemiological parameters may vary from country to country. Confirmed COVID-19 cases in China have been used to estimate those parameters, that vary largely (reviewed in 1). The estimates also depend on testing frequency and case definitions that are prone to change during ongoing epidemics, providing additional uncertainties. The fatal cases due to SARS-CoV2 could be a more reliable parameter, since missing of deaths is less likely. In the absence of changes in the management of severe COVID-19 cases, the rise and decline in virus infections should be followed by a proportional rise and decline in death cases. Although the fluctuating low numbers of fatal cases very early in the epidemic may lead to some uncertainty, more than 100 deaths per day are reported since 10.03.2020 in Italy and since 21.03.2020 in the US. Therefore, the dynamics of new infection and death cases were analysed to estimate the daily reproduction numbers (R_t_) and the effectiveness of control measures in the most affected European Countries and the US.

The daily number of new infection and death cases from 21.2.2020 to 12.04.2020 were downloaded from ECDC (2). A seven-day sliding period was used to smoothen day to day variations. Based on two independent studies we assumed a serial interval of 4 days for SARS-CoV-2 (3, 4).To estimate the daily reproduction number (R_t_) in a simple comprehensible manner, we determined the fold-increase of new infections and death cases during the duration of one serial interval. This way, the fold enhancement during one serial interval can be directly taken as the R_t_ value. Applying this calculation to the daily infection cases in Italy, Spain, France, Germany, UK and the US revealed initial daily reproduction numbers (R_t_) between 2 and 3.55 (Fig. 1). Spontaneous changes in behaviour, implementation of travel restrictions and contact reduction measures probably all contributed to the reduction in R_t_ observed. Calculating R_t_ based on the reported daily death cases also revealed a decline in the R_t_ in each country with a delay of several days. In France and the UK peaks in death based R_t_were not preceded by peaks in R_t_ based on the number of new infections. This may be due to insufficient diagnostic test capacities, limiting the detection of new infections as the numbers of new infections rise steeply. R_t_ based on infections or deaths converged to nearly identical values in all countries analysed. This is expected, if transmission rates remain nearly unchanged for a period longer than the average interval between the time of diagnosis and the time of death. The R_t_ values at the time of convergence are in the range of 1 and seem to level off in countries that have implemented strict contact reduction measures for the longest period of time. A factor impeding a further decrease of R_t_ may be ongoing virus spread in highly susceptible subpopulations such as nursing home residents and highly exposed nursing and medical staff.

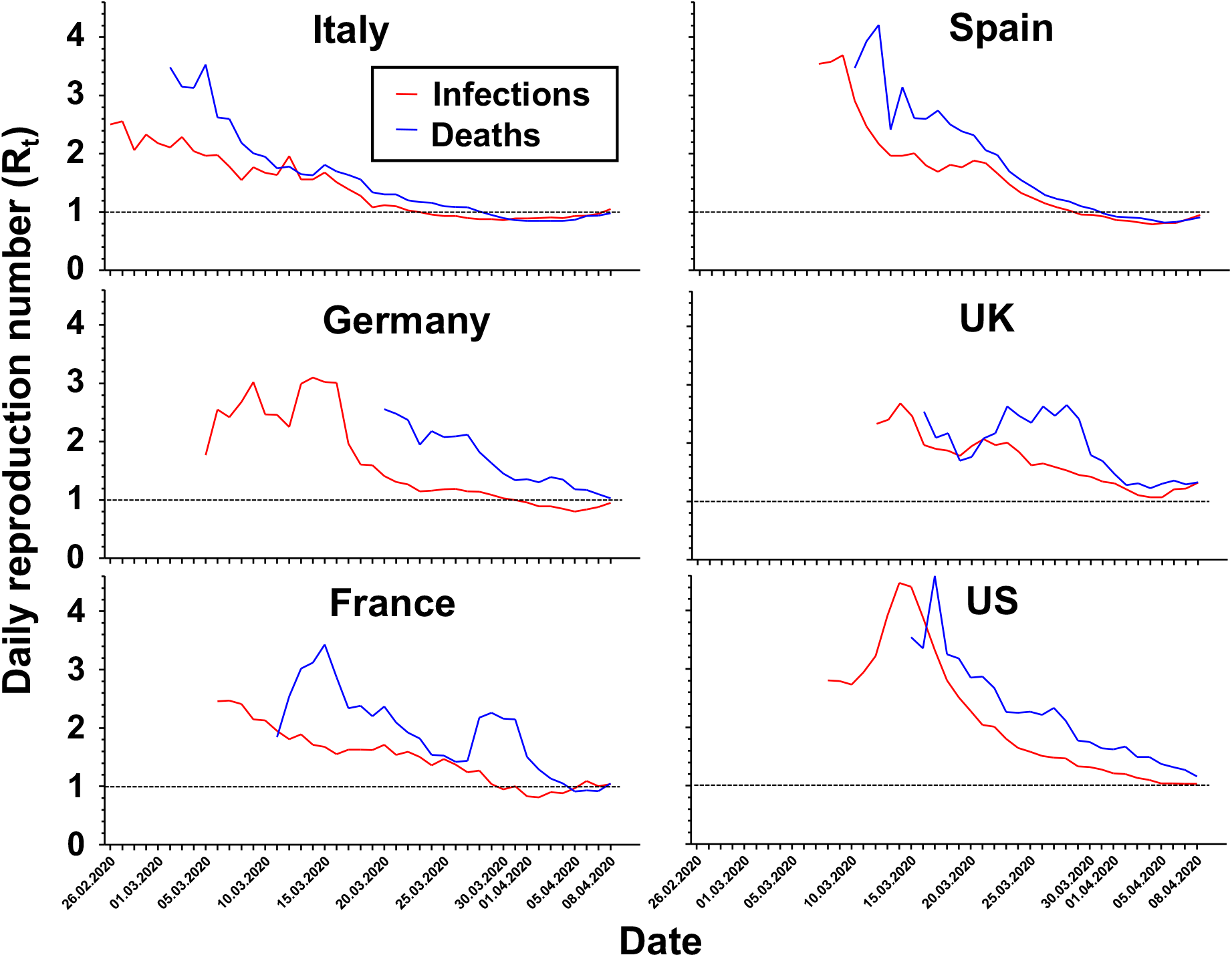
Course of daily reproduction numbers estimated from the number of newly reported SARS-CoV-2 infections or deaths in the indicated countries. To estimate the reproduction numbers, R_t_, we first determined the mean of the number of infections or deaths reported on the indicated day, the three proceeding days and the three subsequent days. The fold-enhancement of the mean number of infections or deaths for each day during the next four days (corresponding to one serial interval) represents the R_t_ estimate shown. Data are presented if the mean number of infections or deaths exceeded 100 and 10 events per day, respectively. Mean number of infections and deaths on the 10^th^, 11^th^ and 12^th^ of April were calculated based on the available 6, 5, and 4 data points, respectively. Virus infection occurs approximately 19 to 29 days before the day of death assuming 4 to 7 days of mean incubation period (5) and 15 to 22 days from onset of symptoms to death (6). Thus, the death based R_t_ values plotted for each day reflect transmissions occurring approximately 24 days before. Similarly, infection based R_t_ values for each day also reflect transmissions occurring several days before. Delays in testing and reporting further limit the accuracy for defining the actual period to which the R_t_ values refer to.

One interpretation of these data is that voluntary changes in behaviour, travel restrictions, and contact reduction measures clearly reduced the R_t_ in all countries analysed to values around 1. Undoubtedly, reduced adherence to contact reduction measures, in populations tired of these measures or lowering these measures, will to lead to a resurgence in new infections. Based on the current estimates of the R_t_ the number of new infections and deaths are expected to decline only slowly, if at all. For countries, in which death based R_t_ have not yet levelled off (e.g. Germany, US), the effectiveness of the national measures implemented to reduce virus spread can currently not be firmly assessed. Despite uncertainty in the reliability of the data used and lack of information on possible changes in the effectiveness of registration of COVID-19 deaths during the observation period, our findings should be considered as a working hypothesis demanding further investigations. As the number of deaths rapidly increases worldwide, we encourage more sophisticated modelling of the epidemic based on the dynamics of death cases by experts in the field. In summary, calculating R_t_ based on the daily number of deaths as well as of new infections may lead to more reliable estimates than those based on infection cases alone, as death based R_t_ are expected to be less susceptible to testing bias or limited capacities.

## Data Availability

Data are publicly available

